# Acute and Post-Acute COVID-19 Outcomes Among Immunologically Naïve Adults During Delta Versus Omicron Waves

**DOI:** 10.1101/2022.11.13.22282222

**Authors:** Margaret K. Doll, Alpana Waghmare, Antje Heit, Brianna Levenson Shakoor, Louise E. Kimball, Nina Ozbek, Rachel L. Blazevic, Larry Mose, Jim Boonyaratanakornkit, Terry L. Stevens-Ayers, Kevin Cornell, Benjamin D. Sheppard, Emma Hampson, Faria Sharmin, Benjamin Goodwin, Jennifer M. Dan, Tom Archie, Terry O’Connor, David Heckerman, Frank Schmitz, Michael Boeckh, Shane Crotty

**Author notes:** these authors contributed equally as co-first authors (Margaret K. Doll, Alpana Waghmare). these authors contributed equally as co-senior authors (Michael Boeckh, Shane Crotty). **Corresponding author:** Margaret K. Doll, PhD, MPH; Albany College of Pharmacy & Health Sciences, 106 New Scotland Ave, HAB121A, Albany, NY 12208 • •.

## Abstract

II.

**Importance:** The U.S. arrival of the Omicron variant led to a rapid increase in SARS-CoV-2 infections. While numerous studies report characteristics of Omicron infections among vaccinated individuals and/or persons with a prior history of infection, comprehensive data describing infections among immunologically naïve adults is lacking.

**Objective:** To examine COVID-19 acute and post-acute clinical outcomes among a well-characterized cohort of unvaccinated and previously uninfected adults who contracted SARS-CoV-2 during the Omicron (BA.1/BA.2) surge, and to compare outcomes with infections that occurred during the Delta wave.

**Design:** A prospective cohort undergoing high-resolution symptom and virologic monitoring between June 2021 and September 2022

**Setting:** Multisite recruitment of community-dwelling adults in 8 U.S. states

**Participants:** Healthy, unvaccinated adults between 30 to 64 years of age without an immunological history of SARS-CoV-2 who were at high-risk of infection were recruited. Participants were followed for up to 48 weeks, submitting regular COVID-19 symptom surveys and nasal swabs for SARS-CoV-2 PCR testing.

**Exposure(s):** Omicron (BA.1/BA.2 lineages) versus Delta SARS-CoV-2 infection, defined as a positive PCR that occurred during a period when the variant represented _≥_50% of circulating SARS-CoV-2 variants in the participant’s geographic region.

**Main Outcome(s) and Measure(s):** The main outcomes examined were the prevalence and severity of acute (_≤_28 days post-onset) and post-acute (_≥_5 weeks post-onset) symptoms.

**Results:** Among 274 immunologically naïve participants, 166 (61%) contracted SARS-CoV-2. Of these, 137 (83%) and 29 (17%) infections occurred during the Omicron- and Delta-predominant periods, respectively. Asymptomatic infections occurred among 6.7% (95% CI: 3.1%, 12.3%) of Omicron cases and 0.0% (95% CI: 0.0%, 11.9%) of Delta cases. Healthcare utilization among Omicron cases was 79% (95% CI: 43%, 92%, *P* =0.001) lower relative to Delta cases. Relative to Delta, Omicron infections also experienced a 56% (95% CI: 26%, 74%, *P* =0.004) and 79% (95% CI: 54%, 91%, *P* <0.001) reduction in the risk and rate of post-acute symptoms, respectively.

**Conclusions and Relevance:** These findings suggest that among previously immunologically naïve adults, few Omicron (BA.1/BA.2) and Delta infections are asymptomatic, and relative to Delta, Omicron infections were less likely to seek healthcare and experience post-acute symptoms.

**KEY POINTS:** *Question:* What are acute and post-acute outcomes among previously uninfected and unvaccinated adults who contracted Omicron (BA.1/BA.2), and how do these compare with Delta infections?

*Findings:* In this prospective cohort of 274 immunologically naïve adults, 166 (61%) contracted SARS-CoV-2, with 9 (5.5%) asymptomatic infections. Compared with Delta, Omicron infections experienced a 79% relative reduction in healthcare utilization, and 56% and 79% relative reductions in the risk and rate of post-acute symptoms (_≥_5-weeks), respectively.

*Meaning:* These findings suggest among immunologically naïve adults, few infections are asymptomatic, and relative to Delta, Omicron infections have lower likelihoods of severe illness and post-acute symptoms.

## Introduction

COVID-19 public health policies and control efforts must consider evolving clinical and epidemiological features of disease.^1-3^ Since these features can be impacted by SARS-CoV-2 genomic changes, the World Health Organization recommends re-evaluation of the clinical course of COVID-19 with the arrival of new SARS-CoV-2 variants.^3^ In December 2021, the B.1.1.529 (Omicron) variant of concern became the predominant SARS-CoV-2 variant circulating in the United States (U.S.), followed by a rapid rise in cases. By March 2022, the U.S. seroprevalence of infection-induced SARS-CoV-2 antibodies was estimated to have increased by nearly 25%,^4^ with a 35% increase observed among unvaccinated adults.^5^

Although numerous studies have found differences in Omicron variant clinical outcomes compared with previous variants,^6-11^ these findings are difficult to interpret due to changes in population immunity from natural infection and/or vaccinations.^12-14^ To date, longitudinal, community-based cohort studies examining Omicron infection characteristics have predominantly studied vaccinated cohorts.^7,15^ Yet, investigating clinical outcomes among immunologically naïve populations remains important to understand the consequences of SARS-CoV-2 genomic variation.

In this study, we examined acute and post-acute (or “long”) COVID-19 clinical outcomes during a period of Omicron (BA.1/BA.2 lineages) variant predominance among a community-based prospective cohort of immunologically naïve adults who were at high-risk of infection and undergoing high-resolution symptom and virologic surveillance. To identify differences in clinical illness, we compared outcomes with study participants who contracted SARS-CoV-2 during a period of Delta variant predominance.

## Methods

### Design, Setting, and Participants

The COVID-19 Immune Protection Study (CovidIPS) is a multisite, prospective cohort study designed to examine SARS-CoV-2 innate and adaptive immune responses among immunologically naïve adults. To ensure an adequate sample size, participants were recruited using web-based advertisements from targeted geographic locations with high SARS-CoV-2 community transmission and suboptimal COVID-19 vaccination rates in 8 U.S. states (AL, AZ, CA, ID, NV, OR, UT, and WA). To ensure feasibility of study procedures, recruitment locations must have been within the service-region of a national mobile phlebotomy company or in close-proximity to one of two research sites.

Volunteers were recruited between March 2021 and February 2022. At enrollment, eligible volunteers were 30 to <65 years of age, without a previous history of COVID-19 vaccination or SARS-CoV-2 infection, and were deemed at high-risk for contracting SARS-CoV-2 (supplementary methods). Volunteers were screened via an electronic survey in English or Spanish. Eligible volunteers were contacted via telephone, with written consent documented electronically. The Western Institutional Review Board approved this study.

### Data and Sample Collection

Study data were collected and managed using REDCap (Research Electronic Data Capture), a secure, web-based software platform designed to support data capture for research studies, hosted by the Fred Hutchinson Cancer Center.^16,17^ To confirm immune status upon enrollment, participants submitted a blood sample, and SARS-CoV-2 receptor binding domain (RBD) IgG endpoint titers were quantified. Participants who had either a positive RBD IgG or a borderline RBD IgG and positive nucleocapsid IgG were excluded from the study. Following enrollment, participants completed a baseline questionnaire, and began weekly “routine” procedures for 24 weeks consisting of both: (i) an electronic survey to report COVID-19 symptoms, and (ii) a self-collected nasal swab submitted for SARS-CoV-2 reverse transcription polymerase chain reactions (RT-PCR) testing (supplementary methods). If a participant reported COVID-19 symptoms or had a RT-PCR-positive swab, the participant was placed on “enhanced” procedures consisting of: (i) daily, electronic symptom surveys and (ii) self-collected nasal swabs submitted every other day, for up to 14-days. Participants who were SARS-CoV-2 positive and reported symptoms (or did not complete their daily symptom survey) on day 14 of enhanced procedures were sent symptom surveys for an additional 14 days (*i*.*e*., through day 28 post-initiation of enhanced procedures).

After 24 weeks, participants completed an end of study blood draw and questionnaire. Participants were also offered an opportunity to reconsent for an additional 24-weeks of follow-up, consisting of the same study procedures as the previous 24-weeks.

### Exposure ascertainment

SARS-CoV-2 infection was defined as _≥_1 RT-PCR-positive result with an infection index date during the time periods that Delta or Omicron (BA.1/BA.2) variants represented the predominant (_≥_50%) SARS-CoV-2 variant circulating in the participant’s geographic region. Weighted, regional NowCast model estimates from the U.S. Centers for Disease Control and Prevention’s national genomic surveillance system were used to ascertain dates to categorize Omicron (BA.1/BA.2) versus Delta infections (supplemental methods).^18^

The infection index date was defined as the first of either the participant’s symptom onset or first SARS-CoV-2 RT-PCR-positive swab collection date. Symptom onset dates were determined by participant self-report; where missing, the first date a symptom was reported on surveys was used.

### Outcome ascertainment

Acute COVID-19 symptoms were defined as symptoms within 28 days of the participant’s symptom onset date, which represents a time period frequently used to demarcate the transition to post-COVID conditions.^19-21^ To minimize misclassification due to nonspecific conditions, a window of ±14 days within the collection date of the first RT-PCR positive result was used to examine COVID-19 symptoms; to be considered asymptomatic, participants must have both: (i) denied symptoms in all surveys submitted during this time period, and (ii) submitted at least one survey indicating they did not have symptoms 7 or more days after RT-PCR testing to ensure sufficient follow up.^2^ Post-acute symptoms were defined as any symptom the participant reported as “related to their previous COVID-19 illness” 5 weeks or more after symptom onset (or first PCR positive for asymptomatic participants); this time period was selected because it was consistent with study procedures (*i*.*e*., a return to weekly procedures at up to 28 days following enhanced procedures), and common definitions of post-COVID conditions as symptoms experienced at least 4 weeks after infection.^19-21^ In sensitivity analyses, we explored alternate post-acute definitions modifying this time period, the number of surveys required with reports of post-acute symptoms, and/or post-acute symptoms considered. Participants recorded post-acute symptoms in routine weekly surveys until the first of either the participant’s study end date or the analyses end date (September 9, 2022).

### Statistical Analyses

Risks and 95% confidence intervals (CI) of acute asymptomatic infections and post-acute symptoms were estimated overall and by variant among participants immunologically naïve at the time of infection; relative risks (*RR*) and 95% CI were estimated comparing Omicron with Delta (referent) infections. Where *RR* could not be estimated (due to zero outcomes among the referent group), absolute risk differences and 95% CI were estimated. To account for differences in post-acute symptom follow up time by variant, rate ratios with 95% CI were estimated using a generalized estimating equation (GEE) approach with a Poisson distribution and offset term to account for completed surveys. In these analyses, a robust “sandwich” covariance estimator with small sample size correction was used to estimate 95% CI using an exchangeable correlation structure.^22,23^ These models were extended to include multivariable adjustment for participant age (continuous) and gender (binary).

Descriptive analyses were performed to analyze the prevalence, mean number, and mean severity of acute or post-acute symptoms in relation to participant symptom onset. Exploratory multivariable analyses were also conducted to examine the relationship between acute illness and post-acute symptoms (supplemental methods).

Baseline and end of study self-rated overall health, memory and concentration, and ability to walk or climb stairs were compared by SARS-CoV-2 variant. For each variable, a change score analysis was performed, comparing the difference in the participant’s end of study response from baseline after adjustment for their baseline score; an alpha level of <0.05 was used to denote statistical significance. In primary analyses, Omicron and Delta infections were directly compared; in secondary analyses, each variant was compared with participants who remained naïve at the end of study survey. For inclusion in either analysis, SARS-CoV-2-positive participants must have had _≥_5 weeks between their end of study survey and symptom onset (or first PCR-positive date for asymptomatic participants).

All analyses were conducted using RStudio with R version 4.2.0 (The R Foundation for Statistical Computing, Vienna, Austria).

## Results

During the study period, 274 immunologically naïve participants were enrolled in the CovidIPS cohort. Among these participants, 166 (61%) became SARS-CoV-2 RT-PCR-positive without a prior history of vaccination or disease; 137 (83%) and 29 (17%) infections occurred during the Omicron- and Delta-predominant periods, respectively. Acute symptom data was available for 164 (99%) infections, post-acute symptom data was available for 150 (90%) infections, and end of study survey data _≥_5-weeks post-infection were available for 133 (80%) participants.

Demographics of the CovidIPS cohort are compared by infection status in Supplemental Results Table 1, and a study flow diagram is provided in Supplemental Figure 1.

**Table 1.** Modified definitions, risks, and risk ratios (*RR*) of post-acute symptoms comparing Omicron versus Delta (referent) variant infections.

| Definition | # Weeks | # Surveys with symptoms | Symptoms | Risk - Omicron (BA.1/BA.2) % (95% CI) | Risk – Delta (B.1.617.2) % (95% CI) | Risk Ratio (95% CI) | P-value |
| --- | --- | --- | --- | --- | --- | --- | --- |
| 1* | ≥5 | ≥1 | Any | 21% (14%, 29%) | 48% (29%, 68%) | 0.4 (0.3, 0.7) | 0.004 |
| 2 | ≥5 | ≥1 | Cognitive<sup>+</sup> only | 12% (7%, 19%) | 15% (4%, 34%) | 0.8 (0.3, 2.3) | 0.7 |
| 3 | ≥5 | ≥2 | Any | 13% (8%, 20%) | 41% (22%, 61%) | 0.3 (0.2, 0.6) | <0.001 |
| 4 | ≥8 | ≥1 | Any | 20% (13%, 28%) | 44% (25%, 65%) | 0.4 (0.3, 0.8) | 0.008 |
| 5 | ≥8 | ≥2 | Any | 8% (4%, 14%) | 37% (19%, 58%) | 0.2 (0.1, 0.5) | <0.001 |
| 6 | ≥12 | ≥1 | Any | 10% (5%, 17%) | 42% (23%, 63%) | 0.2 (0.1, 0.5) | <0.001 |
| 7 | ≥12 | ≥2 | Any | 4% (1%, 9%) | 35% (17%, 56%) | 0.1 (0.0, 0.3) | <0.001 |

<sup>\*</sup>*Definition 1*: used for primary analyses; <sup>+</sup>*Cognitive symptoms* defined as reports of any of the following symptoms: changes in mood, confusion, extreme fatigue, inability to concentrate, insomnia, or memory lapses

### Acute symptoms and healthcare utilization

Among the 164 first-time infections with acute survey data, 9 (5.5%, 95% CI: 2.5%, 10.2%) did not report symptoms and were classified as asymptomatic. Stratified by variant, 0.0% (95% CI: 0.0%, 11.9%) of Delta and 6.7% (95% CI: 3.1%, 12.3%) of Omicron infections were asymptomatic, representing a 6.7% (95% CI: 2.5%, 10.9%, *P* =0.002) absolute risk reduction in symptomatic illness among Omicron infections.

Acute symptom prevalence and severity among the 155 symptomatic infections are examined by days since onset in Figures 1 and 2, while the mean daily number of acute symptoms are examined in Supplemental Figure 2. In general, the prevalence of COVID-19 symptoms was similar by variant; however, participants with Delta infections appeared more likely to report a loss of taste or smell and rate these symptoms with a higher mean severity.

**Figure 1.**
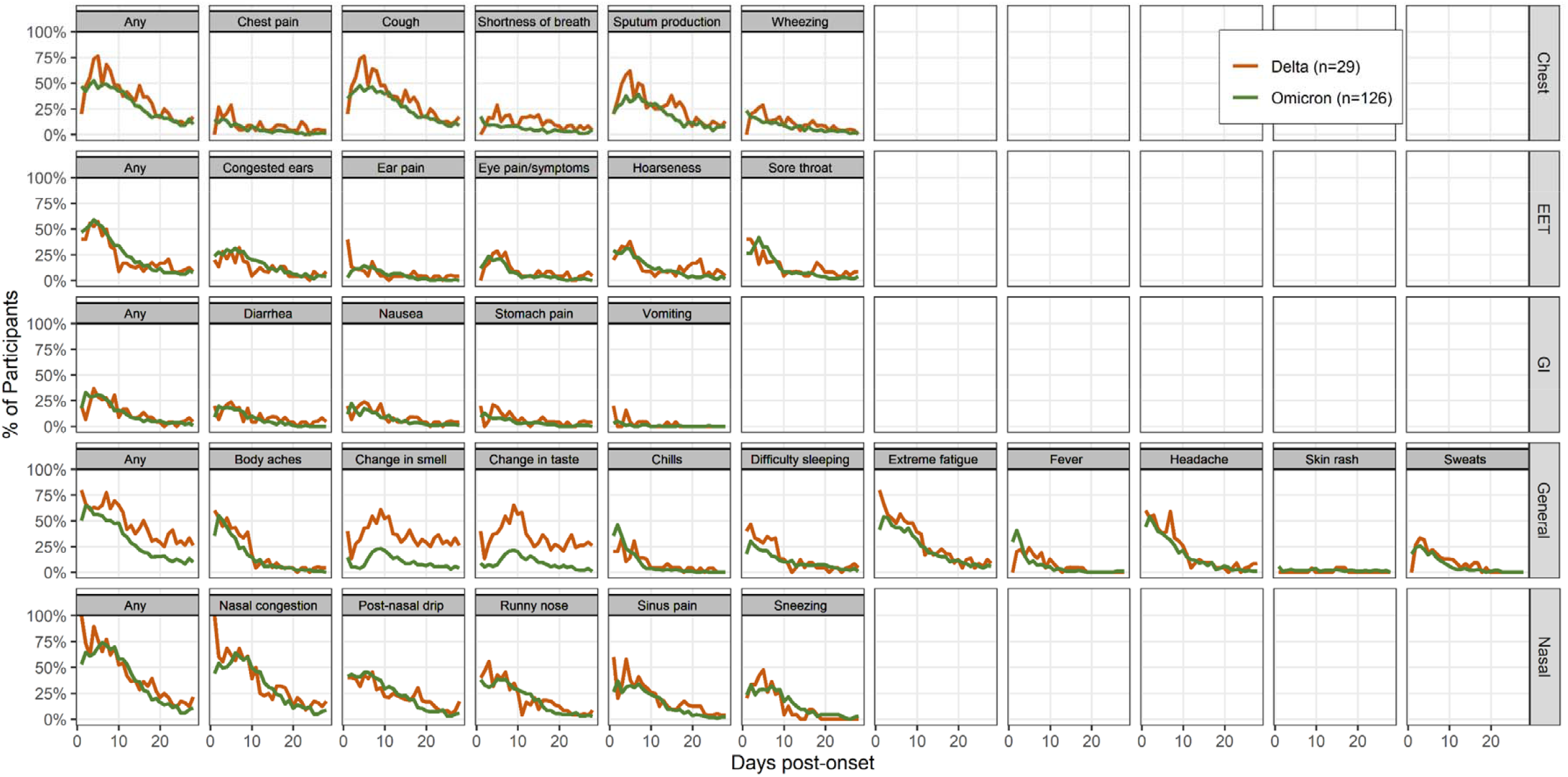
Prevalence of acute COVID-19 symptoms by variant. Data represents the percentage of symptoms reported among participants who submitted a survey each day; only symptomatic persons were included (n=155). *Abbreviations*: EET: eyes, ears, and throat; GI: gastrointestinal.

**Figure 2.**
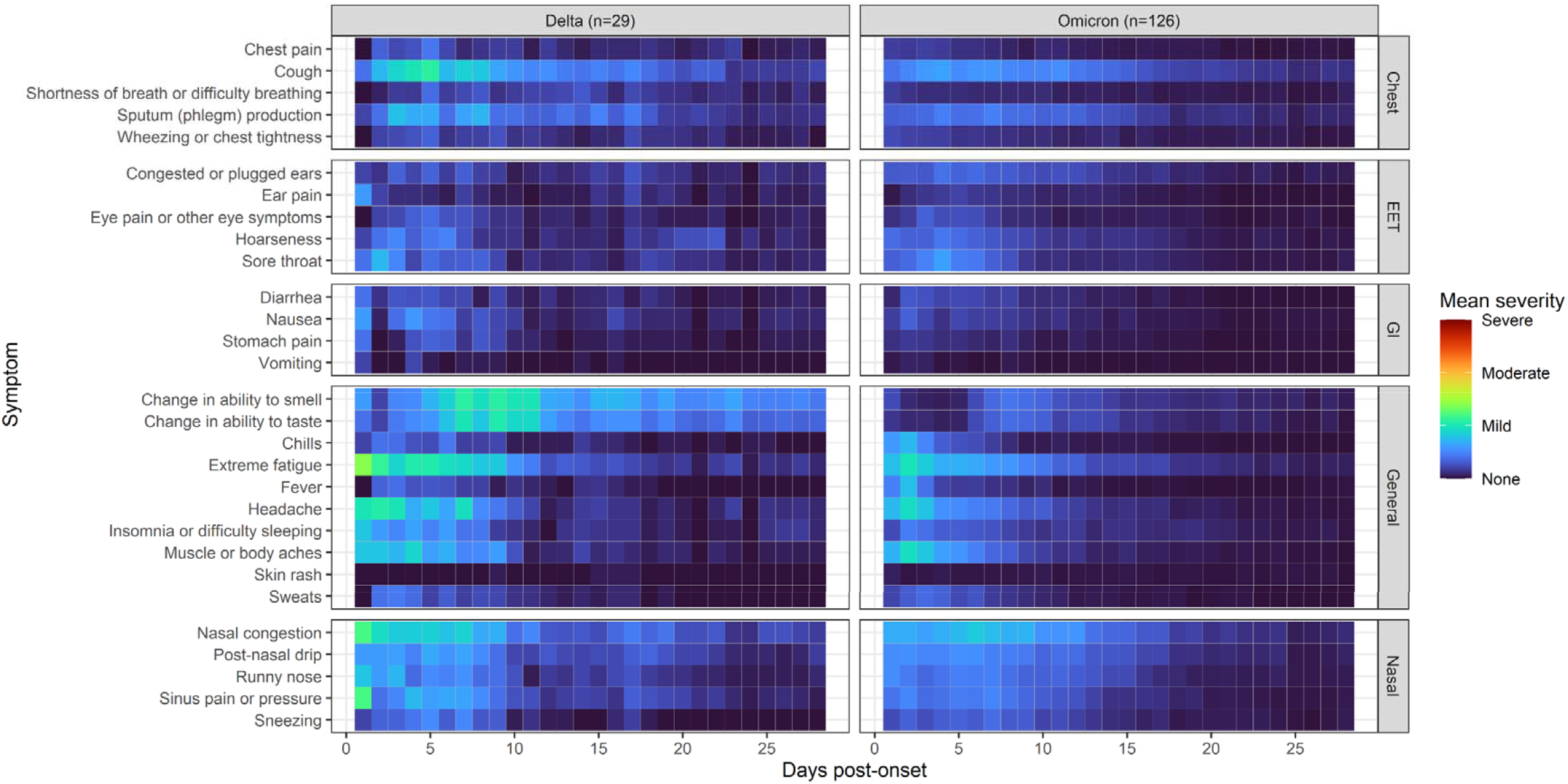
Mean severity of acute COVID-19 symptoms by variant. Data represents the mean severity rating for each symptom among participants who submitted a survey each day; only symptomatic persons were included (n=155). *Abbreviations*: EET: eyes, ears, and throat; GI: gastrointestinal.

Overall, 8.5% (95% CI: 4.7%, 13.9%) of participants sought healthcare for acute symptoms, with no participants requiring emergency care or hospitalization. The *RR* of any healthcare utilization was 79% (95% CI: 43%, 92%) lower among Omicron relative to Delta infections (*RR* = 0.21, 95% CI: 0.08, 0.57, *P*=0.001). Overall, 5.5% (95% CI: 2.5%, 10.2%) of participants received a prescribed medication (supplemental results), with an 83% (95% CI: 40%, 95%, *P*= 0.002) *RR* reduction among Omicron relative to Delta infections.

### Post-acute symptoms

Among 150 participants with post-acute data, 123 and 27 participants were infected during the Omicron- and Delta-predominant periods, respectively. Overall, 39 (26.0%, 95% CI: 19.2%, 33.8%) participants reported at least one symptom during the post-acute period. Stratified by variant, 21.1% (95% CI: 14.3%, 29.4%) of Omicron cases and 48.1% (95% CI: 28.7%, 68.1%) of Delta cases reported post-acute symptoms, representing a 56% (95% CI: 26%, 74%) *RR* reduction in post-acute symptoms (*RR* = 0.44, 95% CI: 0.26, 0.74; *P*=004) among Omicron participants. When analyzed as a rate, Omicron and Delta infections experienced post-acute symptoms 4.9 (95% CI: 3.9, 5.9) per 100 person-weeks and 29.2 (95% CI: 25.4, 33.1) per 100 person-weeks, respectively, representing a rate ratio of 0.21 (95% CI: 0.09, 0.46, *P*<0.001) among Omicron versus Delta infections. Similar rate ratio reductions were found after adjustment for participant age and gender (data not shown). Results from sensitivity analyses demonstrated similar *RR* reductions for most (5/6) modified post-acute definitions (Table 1). In general, the magnitude of *RR* reductions among Omicron participants were more pronounced under more stringent post-acute definitions; however, *RR* estimates were imprecise. A definition examining only cognitive post-acute symptoms found no difference between variants.

We examined mean severity of post-acute symptoms by variant and individual post-acute symptom trajectories in Supplemental Figures 3 and 4, respectively. To ensure comparability, analyses were restricted to 142 (94.7%) unvaccinated participants who did not experience SARS-CoV-2 re-infection; 33 (22 Omicron, 11 Delta) reported post-acute symptoms. While these analyses represent small numbers of participants, they may suggest differences in post-acute symptom severity by strain, with Delta infections reporting higher mean severity and frequencies of post-acute symptoms.

Given the small number of participants with post-acute symptoms, we grouped Delta and Omicron infections together to examine relationships between acute illness and post-acute symptoms. Figure 3 and Supplemental Figures 5 and 6 examine the total number of distinct symptoms, prevalence of acute symptoms, and symptom severity among these participants by post-acute status, while results from exploratory multivariable analyses examining the relationship between acute and post-acute outcomes are reported in Supplemental Results Table 2. In general, these analyses suggest that increasing acute symptom severity, particularly during the first two weeks of acute illness, is associated with higher relative odds/rates of post-acute symptoms, and that prevalence of many acute symptoms was greater among participants who experienced post-acute symptoms.

**Figure 3.**
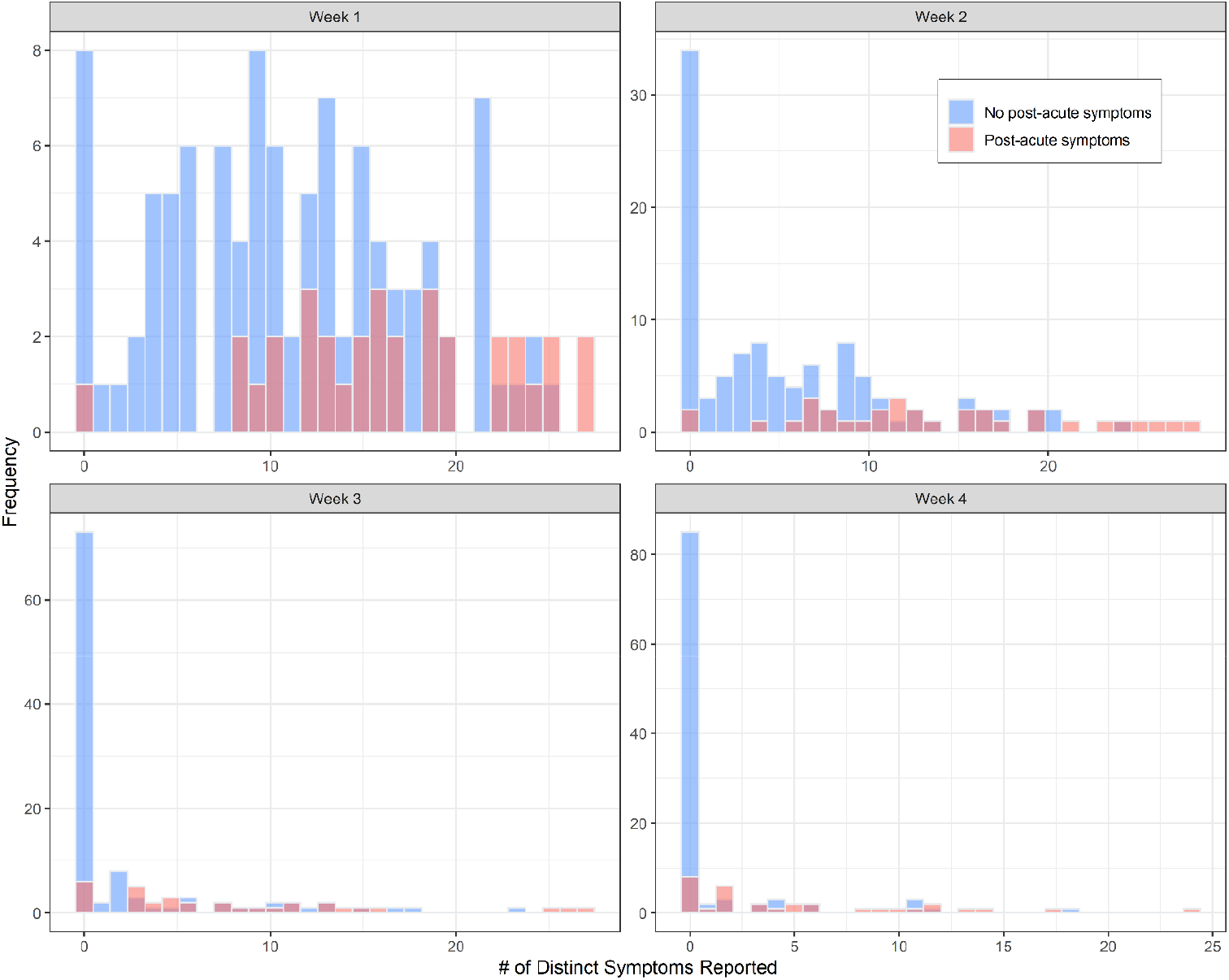
Histogram of distinct acute symptoms reported by week and post-acute symptom status; only unvaccinated and persons with one episode of SARS-CoV-2 were included (n=142).

### Changes in baseline and end of study self-rated scores

Among the 133 (23 Delta, 110 Omicron) participants with end of study data _≥_5-weeks following symptom onset, no changes were detected by variant in self-rated measures for overall health, memory or concentration, or physical abilities between enrollment and end of study surveys (Figure 4). These results were similar for primary analyses comparing variants directly, and secondary analyses comparing each variant to participants who remained naïve at their end of study survey (n=90, Figure 4).

**Figure 4.**
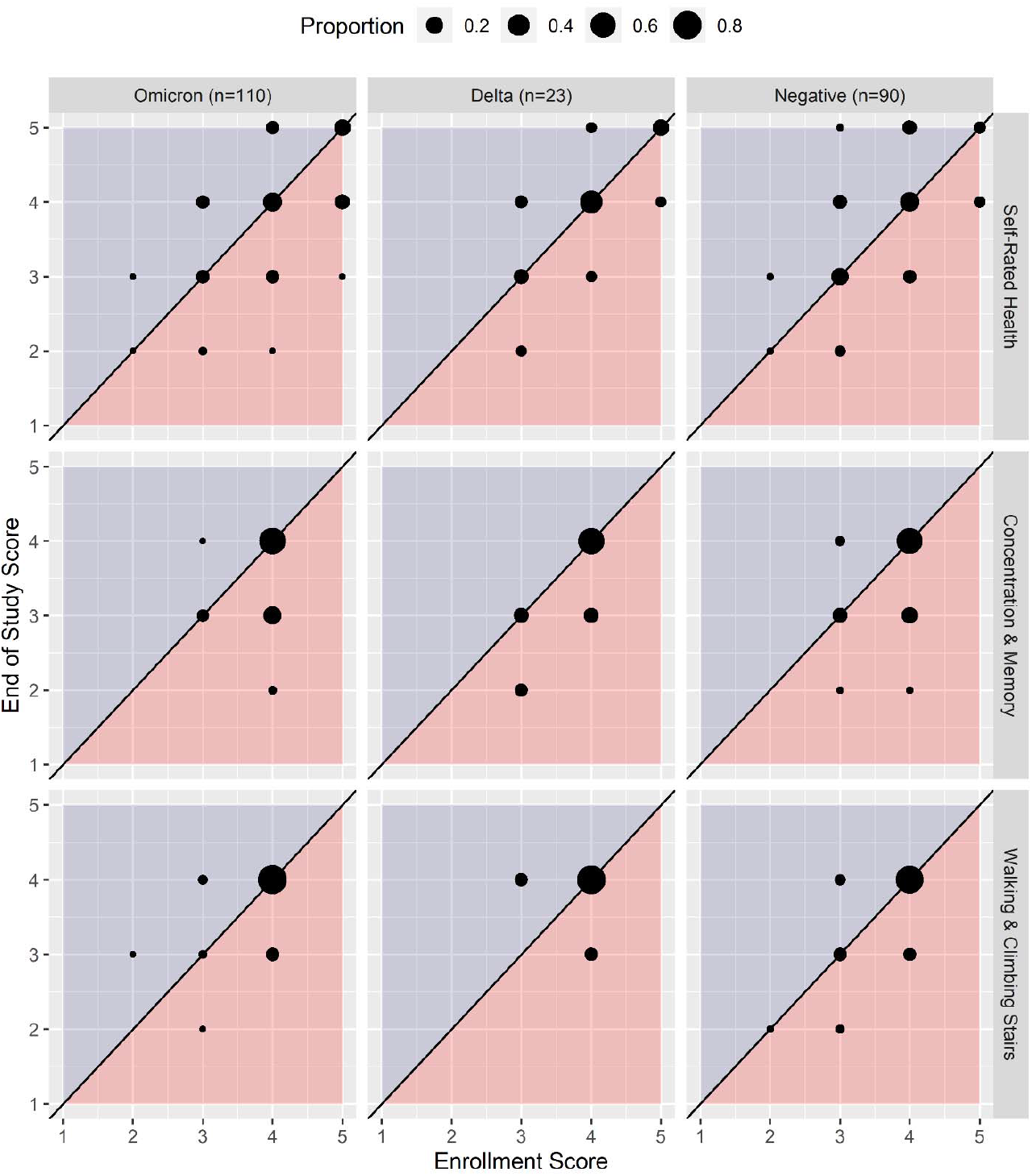
Comparison of participant enrollment and end of study self-rated scores for health (highest health = 5), concentration & memory (highest abilities = 4), and ability to walk & climb stairs (highest abilities = 4). Dots represent the proportion of participants by variant/SARS-CoV-2 status whose responses align with a given survey pattern. Areas shaded in red represent a decline in health scores at the end of study survey, while shaded blue areas represent a positive health gain at the end of study.

## Discussion

In this study, we describe COVID-19 acute and post-acute outcomes in a well-characterized, community-based, prospective cohort of unvaccinated and previously uninfected healthy adults at high risk of infection. Our data demonstrate that SARS-CoV-2 infections during the Omicron (BA.1/BA.2) period were associated with significantly lower absolute risks of acute symptomatic infections, and significant relative reductions in healthcare utilization for acute illness and post-acute symptoms at _≥_5 weeks in comparison with Delta variant infections. While several studies have also found lower acute severity of Omicron compared with Delta infections, these studies have included predominantly vaccinated populations.^7,11^ To our knowledge, this is the first study to characterize both acute and post-acute symptoms in a cohort of immunologically naïve individuals in the era of widespread vaccination and increased SARS-CoV-2 seroprevalence using prospective surveillance.

Defining the spectrum of COVID-19 illness for SARS-CoV-2 variants remains important to elucidate the clinical consequences of SARS-CoV-2 genomic variation.^1-3^ For these purposes, exploring outcomes among unvaccinated and previously uninfected persons may be of particular significance for understanding the natural course of infection in the absence of pre-existing host immunity, facilitating comparisons between variants, and identifying the impact of COVID-19 among non-naïve populations, should immunity wane to levels that do not influence clinical illness.

Importantly, as COVID-19 hospitalization and mortality rates have fallen, other endpoints, such as symptom prevalence and severity, have become critical to characterize the impact of different variants. In the present study, we evaluated epidemiological characteristics of mild COVID-19 illness using a prospective cohort design with frequent and high-resolution symptom and virologic surveillance. With these rigorous methods, we were able to provide key insights regarding the prevalence of asymptomatic infections, which are difficult to ascertain using cross-sectional designs that cannot distinguish between pre-symptomatic or asymptomatic infection,^2^ or cohorts recruited at a healthcare setting, which may be increasingly unrepresentative of mild disease in an era of pervasive at-home SARS-CoV-2 diagnostics.^24^ Overall, we found that the prevalence of asymptomatic infections was only 6% among all SARS-CoV-2 infections, with a significantly higher absolute prevalence among Omicron (7%) versus Delta (0%) infections. Our estimates of asymptomatic Omicron infections were lower when compared with a recent systematic review and meta-analysis, which also found a significantly higher proportion of asymptomatic infections among Omicron (25%) versus Delta (8%) infections.^25^ This apparent difference may reflect characteristics of our study cohort, who represent previously immunologically naïve adults, 30 to 64 years of age, factors also associated with a lower prevalence of asymptomatic disease.^25^

Our prospective design also enabled us to estimate risks of post-acute or “long” COVID-19 symptoms among a fully enumerated cohort of SARS-CoV-2 infections, which may be more accurate than designs that recruit participants at healthcare settings or following the development of post-acute symptoms.^1^ We found that approximately one-quarter (26%) of participants experienced at least one post-acute symptom _≥_5-weeks following their infection. Our study is also among few to date that compared the risks of post-acute COVID-19 symptoms by variant, and the only study (to our knowledge) that compared post-acute risks among unvaccinated individuals. We found that risks of post-acute symptoms at _≥_5 weeks differed significantly by variant, with approximately one-fifth (21%) of Omicron infections versus nearly one-half (48%) of Delta infections experiencing post-acute symptoms, representing a relative risk reduction of more than 50% or a rate ratio reduction of 79% among Omicron infections. These findings remained robust under more stringent definitions of post-acute symptoms examining symptoms at _≥_8- and _≥_12-weeks. Our results were also similar to results from a study of vaccinated individuals, which found 59% to 77% reductions in the odds of post-acute symptoms among Omicron relative to Delta infections.^26^

This study has several strengths. Our recruitment methods and prospective design enabled us to ascertain participant baseline immune status with greater certainty than studies using administrative data. Our study procedures also featured frequent symptom and virologic surveillance with few missing data and high retention of study participants, allowing us to characterize outcomes with greater accuracy than cohorts ascertained following infection. Further, the distribution of SARS-CoV-2 infections across Delta and Omicron time periods permitted us to compare and detect significant differences in variant outcomes using the same study procedures. Finally, during an era of widespread infections and vaccinations, our study population of immunologically naïve adults also represents a unique cohort, who at the time of the study, comprised a significant proportion of the U.S. population, yet remain underrepresented in scientific research.

Our study also has several limitations. We did not sequence viral samples and used U.S. regional prevalence data to classify variants for our SARS-CoV-2 infections. While this approach is similar to methods in previous studies,^7,26^ it is possible that variants were misclassified. Due to the prospective nature of our study and our unique study cohort, our sample size was also smaller in comparison with studies using administrative data.

In conclusion, we demonstrate that among a community-based cohort of immunologically naïve adults, SARS-CoV-2 infections during Omicron (BA.1/BA.2) and Delta predominant periods were associated with few asymptomatic cases of COVID-19 illness, and that Omicron infections were less likely to seek healthcare for acute symptoms and more than half as likely to experience post-acute symptoms relative to Delta infections. Continued study and comparisons of clinical illness from SARS-CoV-2 variants are critical to understand the implications of SARS-CoV-2 genomic variation and must be considered with regard to host immune status.

## Data Availability

Data are not currently available

