## Supplemental Appendix for "Acute and Post-Acute COVID-19 Outcomes Among Immunologically Naïve Adults During Delta Versus Omicron Waves"

**Supplemental Methods**

The following appendix provides additional information regarding the CovidIPS study procedures and statistical methods.

*Participant eligibility – ascertainment of high risk status*

To ensure an adequate sample size of SARS-CoV-2 infections, we developed a scoring system to measure a volunteer’s theoretical risk of contracting SARS-CoV-2. This risk was based upon the following 3 multiple choice survey questions included in the electronic screening questionnaire:

1. How concerned are you about coronavirus (COVID-19)?
2. In an average week, approximately how many people do you have contact with when you are indoors, and NOT wearing a face mask? *Please do not include people who live in your home.*
3. Do you trust that the coronavirus (COVID-19) vaccines are safe?

To be eligible for study participation, volunteers must have received a high-risk score above a pre-determined threshold. Where data from question 3 was not available for a participant, a modified threshold score was applied to ascertain study eligibility.

*Nasal swab collection*

Participants used foam swabs (Custom Swab Puritan Medical Ref #25-18051-PFSC2ARROW) to self-collect a nasal specimen from the mid-nasal space.^27^ Participants were provided written instructions as well as a video link demonstrating sample collection. Participants placed the swab into one nostril and rotated the swabs for 5 rotations. The same swab was then placed in the other nostril also rotated 5 times. The swab was then placed dry into a conical vial and shipped to the lab by the participant. Weekly surveillance swabs were shipped via USPS Priority Mail (1 – 3 days) whereas swabs collected as part of enhanced procedures were shipped via FedEx (week 1 collections: FedEx Priority Overnight; week 2 collections: FedEx 2 Day (3 swabs shipped together at the end of the 2nd week).

*Nasal swab processing*

Upon arrival to the laboratory (Atlas Genomics, Seattle, WA), 2mL of PBS was added to each sample tube and vortexed. Samples were either extracted individually or as pooled samples (Pool of 4: 100uL each sample; Pool of 3: 133uL each sample; Pool of 2: 200uL each sample). RNA extraction was performed using the KingFisher™ Flex Magnetic Particle Processor with 96 Deep-Well Head. 5ul Proteinase K was added to each well along with 200ul of sample. 275ul Binding Bead mix (per well: 265ul Binding Solution, 10ul Total Nucleic Acis Magnetic Beads) was then added followed by 5 μL of MS2 Phage Control. The “MV_Flex” run was used, consisting of a Wash Buffer step, 2 washes with 80% ethanol, and an elution buffer step. The elution plate was either placed on ice for immediate use in real-time RT-PCR or stored at -10 to -30°C freezer for short term storage (1 week).

*RT-PCR testing*

RT-PCR was performed at a diagnostic laboratory (Atlas Genomics Laboratories) based in Seattle, Washington, USA, using the Applied Biosystems™ QuantStudio™ 7 Flex Real-Time PCR Instrument with the TaqPath COVID-19 Combo Kit (Thermo Fisher Scientific).^28^

*SARS-CoV-2 variant exposure classification*

We classified SARS-CoV-2 infections as Omicron **(BA.1/BA.2 lineages)** or Delta variants based upon the participant’s infection index date and the dates these variants represented 50% or more of SARS-CoV-2 variants circulating in the participant’s U.S. Department of Health and Human Services (HHS) geographic region. Participant HHS geographic region was classified according to the mailing address where the participant received their study supplies.

To identify these SARS-CoV-2 variant thresholds, we used weighted, HHS regional **Nowcast model estimates obtained** from **the U.S. Centers for Disease Control and Prevention’s (CDC) national genomic surveillance system.^18^ Calendar dates of SARS-CoV-2 variant circulation by HHS** region are provided in Supplemental Methods Table 1 (below).

**Supplemental Methods – Table 1**

| **HHS Region** | **Participant state address** | **Delta variant (B.1.617.2)**  **Start Date**  **(≥50%; inclusive)** | **Omicron** (**BA.1/BA.2)***  **Start Date**  **(≥50%; inclusive)** | **Omicron** (**BA.1/BA.2)***  **End Date**  **(≥50%; inclusive)** |
| --- | --- | --- | --- | --- |
| **4** | AL | 6/27/2021 | 12/12/2021 | 6/18/2022 |
| **8** | UT | 6/6/2021 | 12/19/2021 | 6/18/2022 |
| **9** | AZ, CA, NV | 6/20/2021 | 12/19/2021 | 6/18/2022 |
| **10** | ID, OR, WA | 6/27/2021 | 12/19/2021 | 6/18/2022 |

**Includes*: B.1.1.529, BA.1.1, BA.2, BA.2.12.1 lineages

*Descriptive analyses examining acute and post-acute symptoms*

For each time unit analyzed in figures, a complete case analysis was performed using data from symptomatic participants who submitted a survey for that time unit, excluding participants with missing data. Participants who reported no symptoms on day 14 were presumed to remain symptom free on days 15-28 following the start of their enhanced procedures. Sensitivity analyses were conducted to exclude infections where symptom onset date may have been misclassified because it was not reported by the participant or observed across surveys.

*Exploratory analyses examining the relationship between acute and post-acute symptoms*

We examined the relationship between acute COVID-19 illness and post-acute symptoms in multivariable analyses using four different modeling approaches to characterize acute and post-acute COVID-19 illness. In these exploratory analyses, we adjusted all models for participant age (continuous, centered at 30 years) and gender (binary), and implemented the following specifications of acute and post-acute symptoms in separate models:

**Acute COVID-19 illness**: Since symptoms of acute COVID-19 vary over the course of illness, we created summary variables to describe acute symptoms for each week of the 4-week duration of the acute illness period. Separate models were then run for weeks 1 to 4 to examine the relationships of distinct weekly patterns with the development of post-acute outcomes. In these analyses, weekly acute COVID-19 symptoms were characterized in two different ways. First, in base models, we expressed weekly acute COVID-19 symptoms as two variables, indicating the: (i) presence/absence of weekly symptoms as a binary variable, and the (ii) total number of distinct weekly acute symptoms experienced to measure symptom severity as a continuous variable. Results of base models informed the approach to characterize acute symptoms in the subsequent models. Where the presence/absence of weekly symptoms was significant ($\alpha<0.05)$, but the total number of symptoms experienced (severity) was not, models were run to explore the presence/absence of 5 specific categories of acute COVID-19 symptoms (Supplemental Methods – Table 2), using data only from persons who experienced symptomatic illness during that week. Where the presence/absence of weekly symptoms was non-significant, but the total number of symptoms (severity) was significant ($\alpha<0.05)$, the 5 categories of symptoms were entered into the model in addition to the term for the total number of symptoms (severity). Where neither term was significant, the 5 specific categories of acute COVID-19 symptoms were entered, and data from all persons (regardless of symptomatic illness) were explored.

**Post-acute symptoms.** In separate analyses, post-acute symptoms ≥5-weeks following onset were examined as either: (i) a dichotomous outcome, representing the presence or absence of symptoms at ≥5-weeks post-onset, or a (ii) a count of the number of study weeks that a participant experienced post-acute symptoms ≥5-weeks following onset. Where post-acute symptoms were expressed as a binary variable (i), we implemented logistic regressions using a generalized linear model (GLM) approach to examine the odds of post-acute symptoms ≥5-weeks following onset. Where post-acute symptoms were expressed as a count, we implemented a generalized estimating equation (GEE) approach with a Poisson distribution and offset term to examine rates of weekly post-acute symptoms.

**Supplemental Methods – Table 2**. Acute symptoms by symptom category.

| **Category** | **Symptom** |
| --- | --- |
| **Nasal** | Nasal congestion |
|  | Post-nasal drip |
|  | Runny nose |
|  | Sinus pain or pressure |
|  | Sneezing |
| **Chest** | Chest pain |
|  | Cough |
|  | Shortness of breath or difficulty breathing |
|  | Sputum (phlegm) production |
|  | Wheezing or chest tightness |
| **General** | Change in ability to smell |
|  | Change in ability to taste |
|  | Chills |
|  | Extreme fatigue (tiredness) |
|  | Fever |
|  | Headache |
|  | Insomnia or difficulty sleeping |
|  | Muscle or body aches |
|  | Skin rash |
|  | Sweats |
| **Eyes, Ears, & Throat (EET)** | Congested or plugged ears |
|  | Ear pain |
|  | Eye pain or other eye symptoms |
|  | Hoarseness |
|  | Sore Throat |
| **Gastrointestinal (GI)** | Diarrhea |
|  | Nausea |
|  | Stomach pain |
|  | Vomiting |

**Supplemental Results**

*Prescription medication use reported for acute COVID-19 illness*

Among participants who had acute symptom data and were infected with Omicron, 4 participants (3.0%, 95% CI: 0.8%, 7.4%) reported receiving a medication for their illness. Medications reported were an antibiotic (n=1), an oral steroid (n=1), ivermectin (n=1), and oseltamivir (n=1). Among participants who had acute symptom data and were infected with Delta, 5 participants (17.2%, 5.8%, 35.8%) reported receiving a medication. Medications reported were an antibiotic (n=2), ivermectin (n=2), and an antibody infusion (n=1).

*Potential misclassification of symptom onset date*

Among participants with acute data (n=164), 14 (2 Delta & 12 Omicron) participants had potentially misclassified symptom onset dates. Exclusion of these cases from analyses which examined the time since symptom onset did not reveal meaningful differences in study results (data not shown).

*Exploratory analyses examining the relationship between acute and post-acute symptoms*

Results from multivariable analyses exploring the relationship between acute and post-acute symptoms are reported in the Supplemental Results - Table 2. In general, we observed different patterns in acute weeks 1 and 2 in comparison with acute weeks 3 and 4. Specifically, we found that the total number of symptoms (severity) in acute weeks 1 and 2 was significantly associated with an increase in the odds/rates of post-acute symptoms. However, in weeks 3 and 4, this pattern was not apparent. For these weeks, the weekly presence/absence of all acute symptoms was instead significantly associated with an increase in the odds/rates of post-acute symptoms. Furthermore, in secondary models with symptom categories, we observed that the presence of chest symptoms in weeks 1 or 2 was (in most cases) significantly/borderline significantly associated with a 4 to 8 times increase in the odds/rates of post-acute symptoms. This was in contrast to weeks 3 or 4, where chest symptoms was non-significant, and instead, the presence of general symptoms was (in some cases) significantly/borderline significantly associated with an increase in the odds/rates of post-acute symptoms. While models comparing symptom categories were largely imprecise, point estimates and trends in results from logistic GLM and Poisson GEE regressions were similar.

**Supplemental Results - Table 1.** Selected demographic and other characteristics among the CovidIPS cohort by SARS-CoV-2 status; note SARS-CoV-2 negatives represent participants who contributed person-time to the study, and did not contract SARS-CoV-2 while immunologically naïve

| **Demographic Variable or Characteristic** | **SARS-CoV-2 Infection: Omicron period**  **(N = 137)** | | **SARS-CoV-2 Infection: Delta period**  **(N = 29)** | | **SARS-CoV-2 Negatives**  **(N = 108)** | |
| --- | --- | --- | --- | --- | --- | --- |
|  | **N** | **% (95% CI)** | **N** | **% (95% CI)** | **N** | **% (95% CI)** |
| **Gender:** Female | 95 | 69 (61, 77) | 20 | 69 (52, 86) | 71 | 66 (57, 75) |
| **Age (years):** Mean | 48 | (47, 50) | 46 | (42, 49) | 51 | (49, 53) |
| **Race (self-report)** |  |  |  |  |  |  |
| White | 127 | 93 (88, 97) | 25 | 86 (74, 99) | 90 | 83 (76, 90) |
| Another | 10 | 7 (3, 12) | 4 | 14 (1, 26) | 18 | 17 (10, 24) |
| **Ethnicity (self-report)*** |  |  |  |  |  |  |
| Not Hispanic or Latino | 122 | 89 (85, 94) | 26 | 90 (83, 100) | 97 | 90 (85, 95) |
| Hispanic or Latino | 10 | 7 (3, 12) | 2 | 7 (0, 18) | 7 | 7 (2, 12) |
| **Education*** |  |  |  |  |  |  |
| High school or equivalent (GED) | 44 | 32 (23, 40) | 2 | 7 (0, 26) | 29 | 27 (18, 38) |
| Associate or technical degree | 38 | 28 (19, 37) | 8 | 28 (10, 47) | 33 | 31 (22, 42) |
| Bachelor’s degree | 35 | 26 (17, 35) | 13 | 44 (28, 64) | 29 | 26 (18, 38) |
| Graduate degree | 19 | 14 (5, 22) | 6 | 21 (3, 40) | 15 | 14 (5, 25) |
| **Employment Status** |  |  |  |  |  |  |
| Not employed | 26 | 19 (10, 28) | 6 | 21 (3, 41) | 30 | 28 (19, 38) |
| Part-time | 49 | 36 (27, 45) | 12 | 41 (24, 61) | 21 | 19 (10, 30) |
| Full time | 62 | 45 (37, 54) | 11 | 38 (21, 58) | 57 | 53 (44, 63) |
| **Occupation** |  |  |  |  |  |  |
| Agriculture, construction, transportation, and manufacturing | 19 | 17 (9, 27) | 2 | 9 (0, 31) | 15 | 19 (9, 31) |
| Arts, entertainment, recreation, retail and wholesale trade | 14 | 12 (7, 29) | 2 | 9 (0, 31) | 5 | 7 (0, 18) |
| Finance, information,  Professional, scientific, management, and public administration | 17 | 16 (11, 32) | 2 | 9 (0, 31) | 14 | 18 (8, 30) |
| Educational services and social assistance | 11 | 10 (4, 25) | 3 | 13 (0, 35) | 4 | 5 (0, 17) |
| Health care | 20 | 18 (10, 28) | 8 | 34 (17, 57) | 18 | 23 (13, 35) |
| Others | 30 | 27 (16, 34) | 6 | 26 (9, 48) | 22 | 28 (18, 403 |
| **Household composition** |  |  |  |  |  |  |
| Living alone | 14 | 10 (2, 18) | 1 | 3 (0, 24) | 19 | 18 (8, 28) |
| 2 persons | 25 | 19 (10 27) | 5 | 17 (3, 38) | 36 | 32 (24, 43) |
| 3 persons | 22 | 16 (8, 24) | 6 | 21 (7, 41) | 20 | 19 (9, 29) |
| 4+ persons | 76 | 55 (47, 64) | 17 | 59 (45, 79) | 33 | 31 (22, 42) |
| **Underlying conditions at baseline** |  |  |  |  |  |  |
| Anxiety | 21 | 15 (9, 21) | 3 | 10 (0, 21) | 22 | 21 (13, 28) |
| Asthma | 10 | 7 (3, 11) | 4 | 14 (1, 26) | 11 | 10 (5, 16) |
| Chronic obstructive pulmonary diseases (COPD) or heart problems | 2 | 1 (0, 3) | 0 | 0 | 9 | 9 (3, 14) |
| Depression | 8 | 6 (2, 10) | 2 | 7 (0, 16) | 10 | 9 (4, 15) |
| Diabetes | 9 | 7 (2, 11) | 2 | 7 (0, 16) | 8 | 8 (2, 12) |
| High blood pressure | 17 | 12 (7, 18) | 3 | 10 (0, 21) | 18 | 17 (10, 24) |
| Post-traumatic stress disorder | 5 | 4 (0, 7) | 0 | 0 | 5 | 5 (0, 9) |
| Thyroid conditions | 11 | 8 (3, 12) | 3 | 10 (0, 21) | 12 | 11 (5, 17) |
| Other medical conditions | 12 | 9 (4, 13) | 5 | 17 (3, 31) | 18 | 17 (10, 24) |
| **Masking behavior** |  |  |  |  |  |  |
| Always | 12 | 9 (0, 17) | 3 | 10 (0, 28) | 18 | 17 (7, 27) |
| Mostly | 36 | 26 (17, 35) | 4 | 14 (0, 31) | 29 | 27 (18, 37) |
| Sometimes | 42 | 30 (22, 40) | 12 | 42 (24, 59) | 36 | 33 (24, 43) |
| Rarely | 39 | 29 (20, 37) | 5 | 17 (0, 35) | 19 | 17 (8, 28) |
| Never | 8 | 6 (0, 14) | 5 | 17 (0, 36) | 6 | 6 (0, 16) |
| **Attends indoor social gatherings** |  |  |  |  |  |  |
| Never | 12 | 9 (0, 18) | 2 | 7 (0, 27) | 19 | 18 (8, 28) |
| Sometimes (≥1x per month) | 53 | 39 (32, 49) | 9 | 31 (17, 51) | 49 | 44 (36, 56) |
| Frequently (≥1x per week) | 72 | 52 (44, 61) | 18 | 62 (48, 82) | 40 | 38 (28, 48) |

*excludes “prefer not to say” category, does not sum to total

**Supplemental Results - Table 2.** Results from multivariable regressions examining the relationship between post-acute symptoms (outcome) and acute COVID-19 symptoms. All analyses are adjusted for gender and age.

| **Model** | **Variable** | **Week 1** | | | **Week 2** | | | **Week 3** | | | **Week 4** | | |
| --- | --- | --- | --- | --- | --- | --- | --- | --- | --- | --- | --- | --- | --- |
| **Logistic GLM** | | **OR** | **95% CI** | **P-value** | **OR** | **95% CI** | **P-value** | **OR** | **95% CI** | **P-value** | **OR** | **95% CI** | **P-value** |
| 1 | Any symptoms | 0.4 | (0.0, 8.9) | 0.459 | 1.2 | (0.2, 9.1) | 0.864 | 6.1 | (1.8, 21.9) | 0.004* | 12.4 | (3.5, 46.3) | <0.001* |
| 1 | Symptom severity | 1.2 | (1.1, 1.3) | <0.001* | 1.2 | (1.1, 1.3) | <0.001* | 1.1 | (1.0, 1.2) | 0.164 | 1.0 | (0.9, 1.2) | 0.495 |
| 2 | Symptom severity | 1.2 | (1.1, 1.4) | 0.008* | 1.2 | (1.1, 1.4) | 0.005* | ---- | | | ---- | | |
| 2 | Nasal symptoms | 0.7 | (0.1, 4.2) | 0.645 | 0.5 | (0.1, 3.6) | 0.503 | 1.4 | (0.2, 10.0) | 0.748 | 0.4 | (0.1, 2.8) | 0.397 |
| 2 | Chest symptoms | 5.8 | (1.3, 34.2) | 0.031* | 8.2 | (1.7, 53.6) | 0.014* | 2.6 | (0.6, 12.7) | 0.209 | 2.5 | (0.4, 20.5) | 0.361 |
| 2 | General symptoms | 1.3 | (0.2, 10.5) | 0.793 | 0.8 | (0.2, 4.2) | 0.778 | 5.3 | (1.3, 27.2) | 0.030* | 4.6 | (1.0, 23.5) | 0.051 |
| 2 | EET symptoms | 0.3 | (0.1, 1.3) | 0.111 | 0.4 | (0.1, 1.4) | 0.160 | 0.8 | (0.2, 3.1) | 0.725 | 1.3 | (0.2, 7.3) | 0.765 |
| 2 | GI symptoms | 0.3 | (0.1, 1.1) | 0.073 | 0.5 | (0.1, 1.7) | 0.268 | 0.3 | (0.1, 1.6) | 0.170 | 0.2 | (0.0, 1.5) | 0.130 |
| **Poisson GEE** | | **RR** | **95% CI** | **P-value** | **RR** | **95% CI** | **P-value** | **RR** | **95% CI** | **P-value** | **RR** | **95% CI** | **P-value** |
| 3 | Any symptoms | 1.0 | (0.1, 14.8) | 0.982 | 2.1 | (0.4, 10.3) | 0.373 | 4.0 | (1.2, 14.1) | 0.028* | 7.9 | (2.6, 24.5) | 0.001* |
| 3 | Symptom severity | 1.1 | (1.1, 1.2) | 0.001* | 1.1 | (1.1, 1.2) | <0.001* | 1.1 | (1.0, 1.1) | 0.107 | 1.1 | (0.9, 1.2) | 0.232 |
| 4 | Symptom severity | 1.1 | (1.0, 1.3) | 0.092 | 1.1 | (1.0, 1.3) | 0.059 | ---- | | | ---- | | |
| 4 | Nasal symptoms | 1.2 | (0.1, 9.2) | 0.88 | 1.5 | (0.3, 7.5) | 0.613 | 0.5 | (0.1, 2.5) | 0.323 | 0.5 | (0.2, 1.5) | 0.169 |
| 4 | Chest symptoms | 4.2 | (1.0, 17.5) | 0.052 | 4.0 | (0.7, 23.5) | 0.126 | 1.8 | (0.5, 6.7) | 0.365 | 1.6 | (0.4, 6.7) | 0.491 |
| 4 | General symptoms | 2.5 | (0.4, 16.5) | 0.314 | 0.4 | (0.1, 2.5) | 0.323 | 5.0 | (0.8, 30.8) | 0.081 | 2.4 | (0.8, 7.6) | 0.123 |
| 4 | EET symptoms | 0.5 | (0.1, 3.0) | 0.400 | 1.1 | (0.3, 4.1) | 0.920 | 1.7 | (0.5, 5.8) | 0.370 | 1.5 | (0.5, 4.3) | 0.434 |
| 4 | GI symptoms | 0.7 | (0.2, 2.2) | 0.517 | 0.7 | (0.2, 2.5) | 0.561 | 0.5 | (0.1, 1.8) | 0.247 | 1.1 | (0.3, 4.5) | 0.887 |

*Denotes statistical significance, based on an alpha level = 0.05.

**Supplemental Figure 1.** Flow diagram detailing enrollment and outcomes in the CovidIPS cohort.

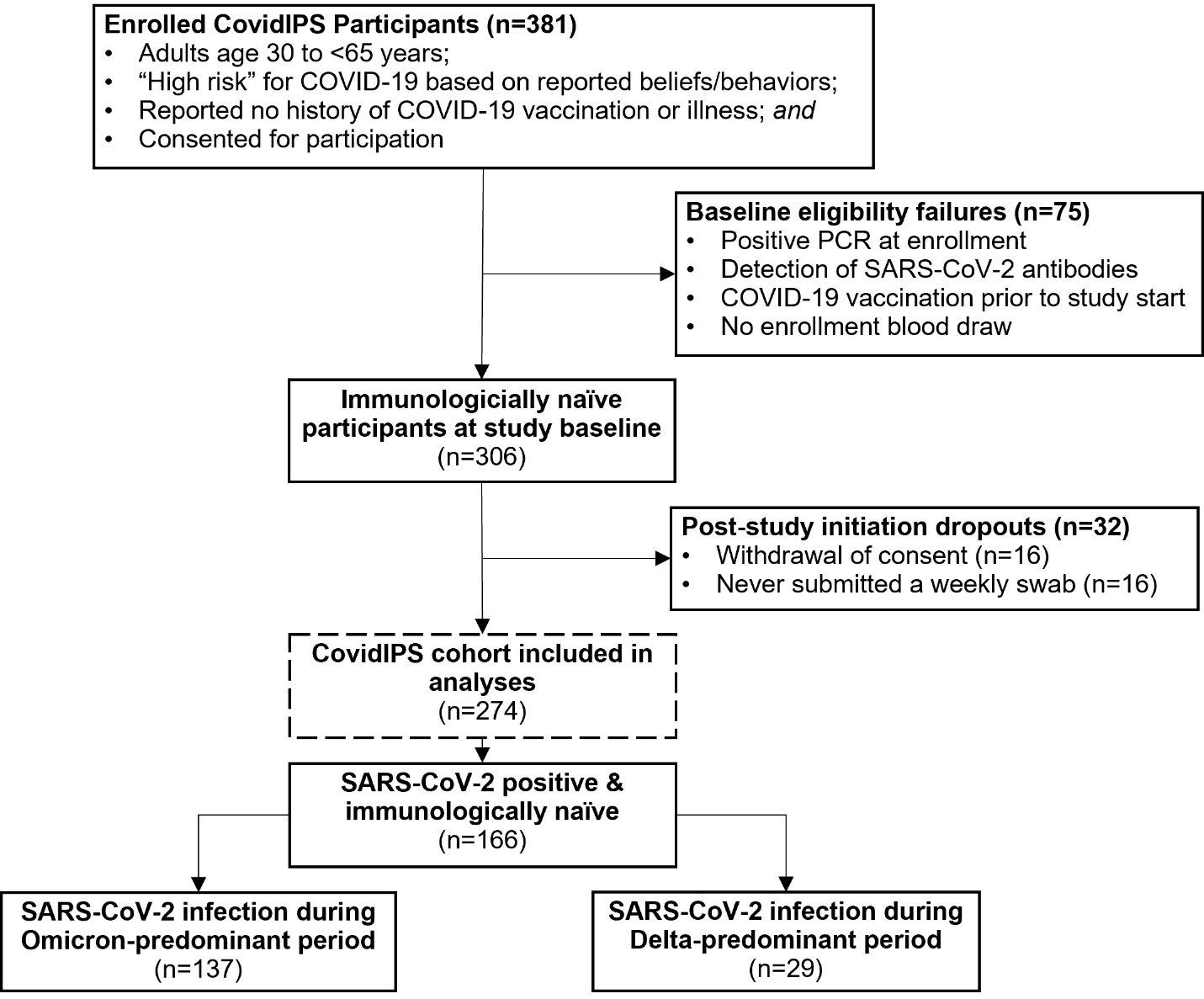

**Supplemental Figure 2.** Mean number of acute symptoms by day and variant. Data represents the mean severity rating for each symptom among participants who submitted a survey each day; only symptomatic persons were included (n=155).

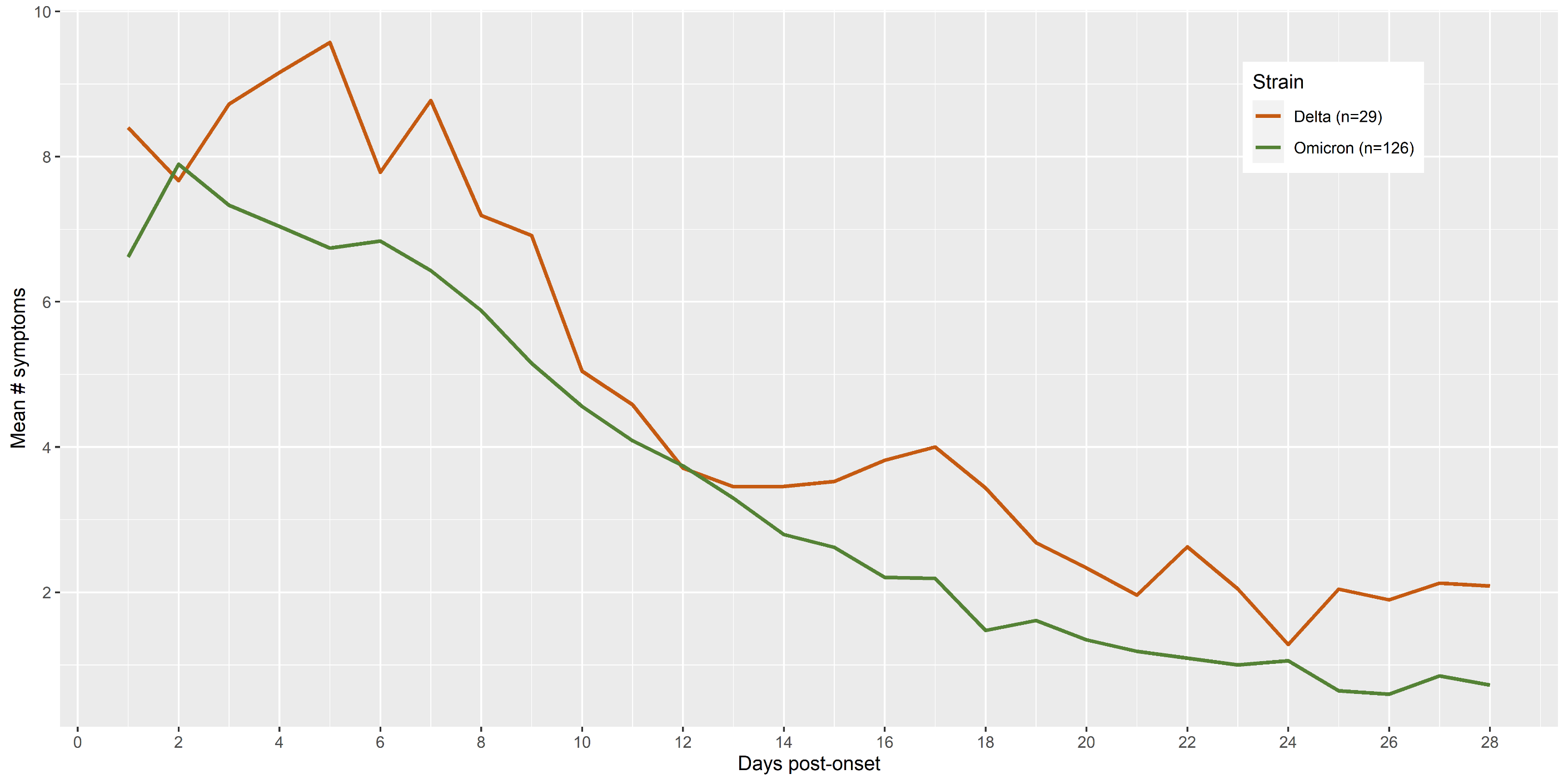

**Supplemental Figure 3.** Mean severity of post-acute COVID-19 symptoms by variant. Data represents the mean severity rating for each symptom among participants who submitted a survey each week; only persons who ever reported any post-acute symptoms were included (n=33). *Abbreviations*: EET: eyes, ears, and throat; GI: gastrointestinal.

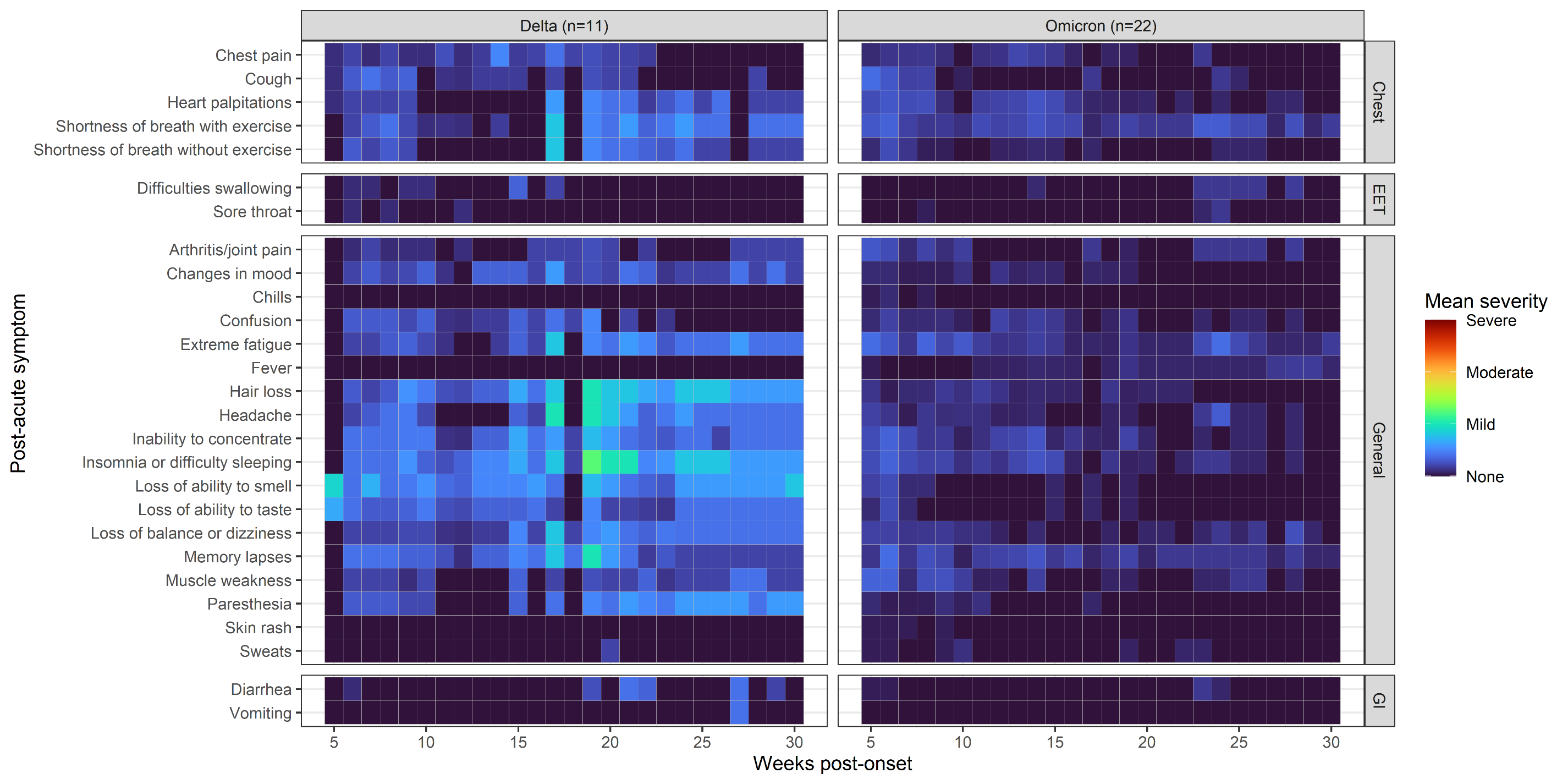

**Supplemental Figure 4.** Trajectory of post-acute COVID-19 symptoms by variant. Each box represents the total number of symptoms reported (max 28) by participants who ever reported post-acute symptoms (n=33) during the post-acute period (≥5 weeks). Shaded grey values represent weeks the participant did not contribute data because the time period was outside the study window (or between primary/extension periods); while missing data due to survey non-response is represented by black “X”s.

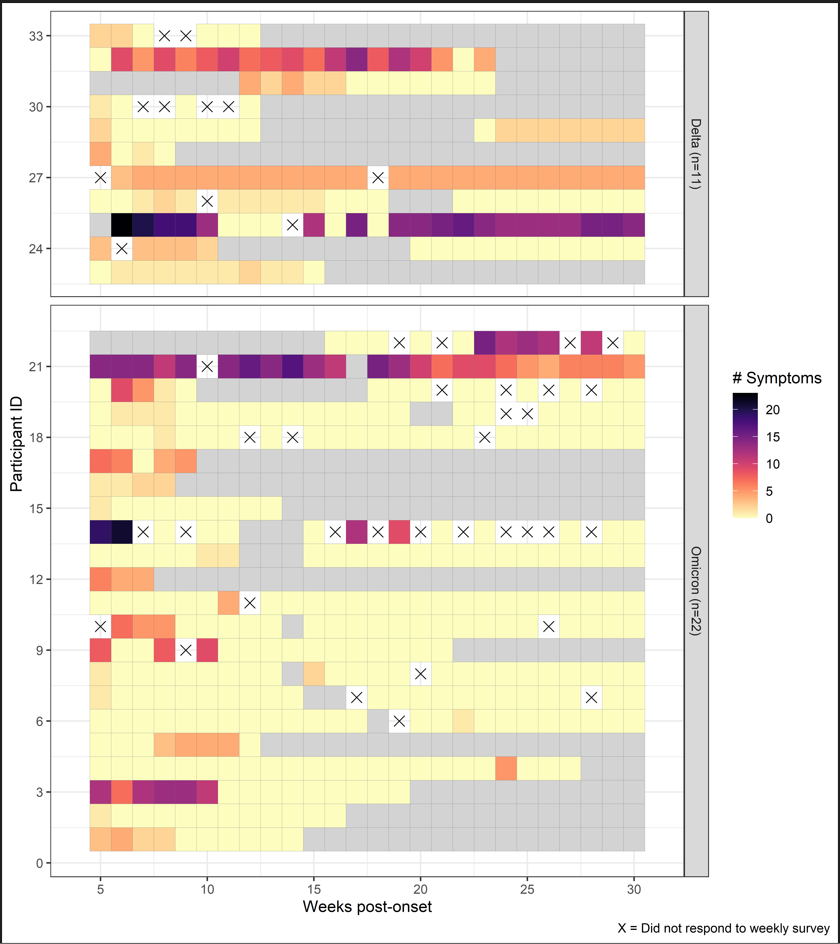

**Supplemental Figure 5.** Prevalence of acute COVID-19 symptoms by post-acute status. Data represents the percentage of symptoms reported among participants who submitted a survey each day; only unvaccinated and persons with one episode of SARS-CoV-2 were included (n=142). *Abbreviations*: EET: eyes, ears, and throat; GI: gastrointestinal.

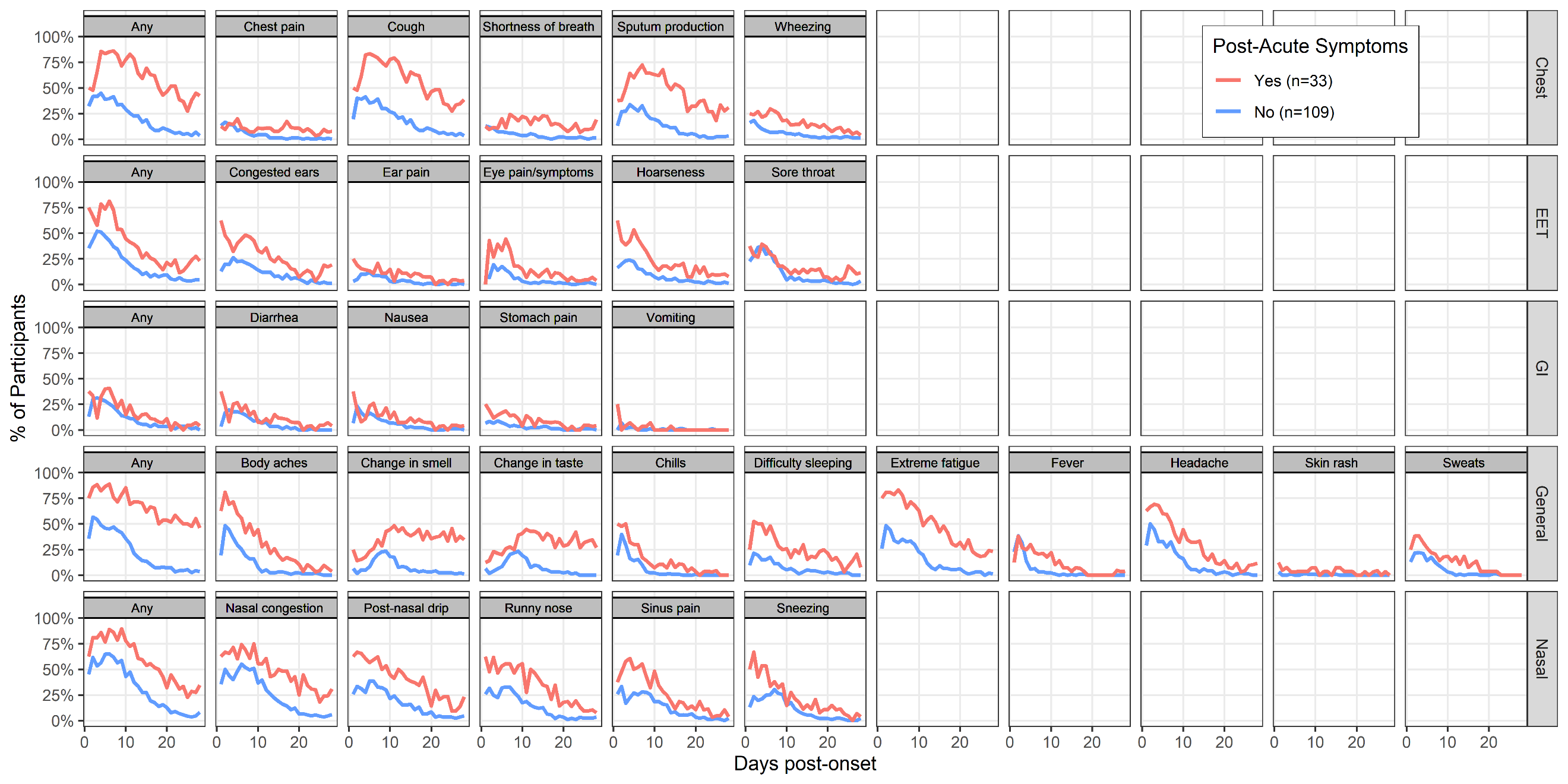

**Supplemental Figure 6.** Mean severity of acute symptoms by day and post-acute status. Data represents the mean severity rating for each symptom among participants who submitted a survey each day; only unvaccinated and persons with one episode of SARS-CoV-2 were included (n=142). *Abbreviations*: EET: eyes, ears, and throat; GI: gastrointestinal.

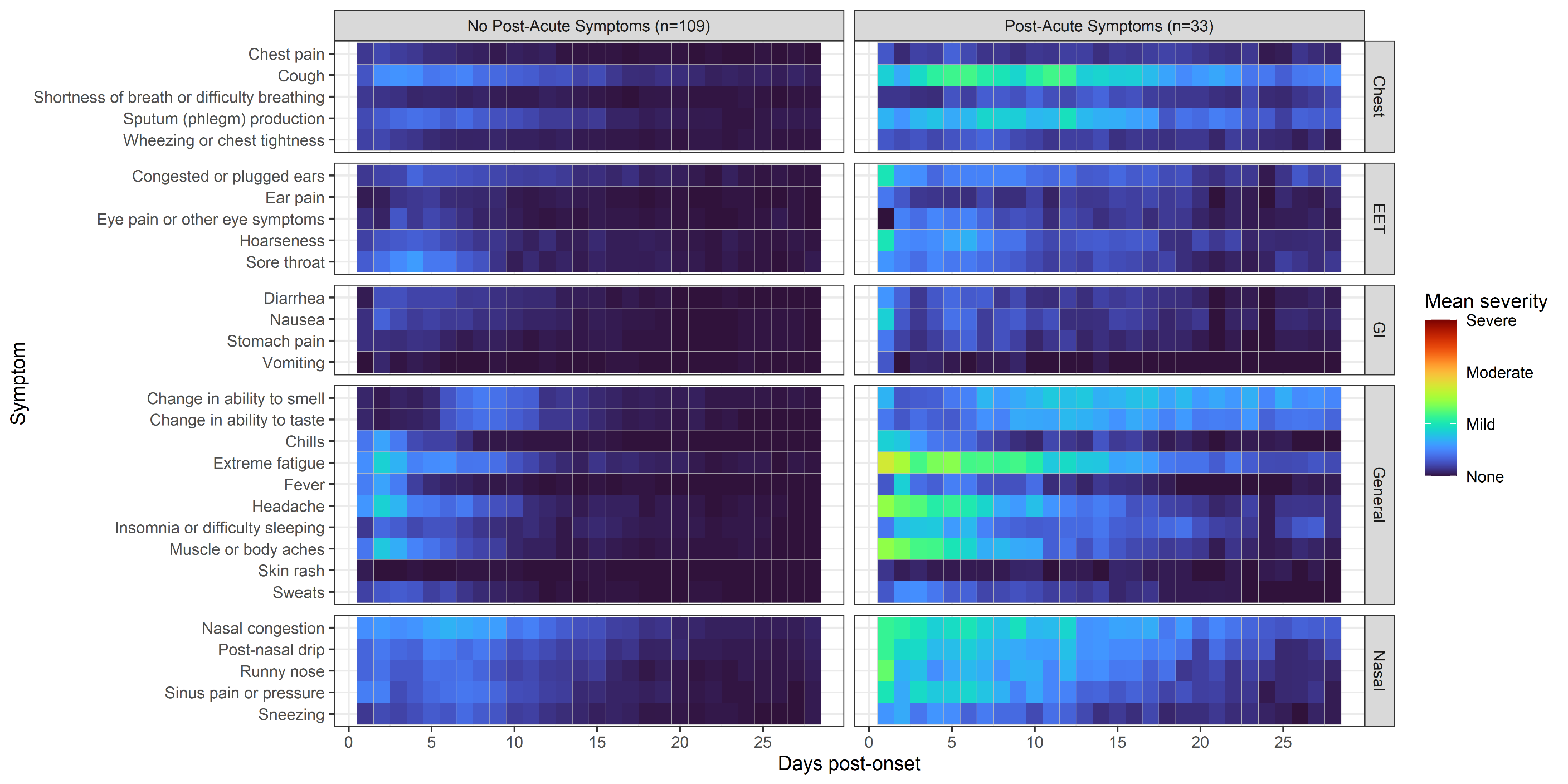
